## Supplementary material for "Physical activity in adult users of inpatient mental health services: a scoping review": S1 File

**S1 File** Search strategy

Mental health terms: **title and abstract**

Mental health or Mental well-being or Mental well being or Psychological well being or Psychological well-being or Mental disorder* or Mental illness* or Mental disease* or Anxiety disorder* or Delirium or Dissociative disorder* or Factitious disorder* or Mood disorder* or Affective disorder* or Depressive disorder* or Neurotic disorder* or Personality disorder* or Schizophreni* or Somatoform disorder* or Adjustment disorder* or Neuros?s or Psychos?s or Delusion* or Paranoia or Hallucination* or Depression or Panic disorder* or Phobia* or Health anxiet* or Bipolar disorder* or Obsessive compulsive disorder* or Obsessive thought* or Intrusive thought* or Post traumatic stress disorder* or Post-traumatic stress disorder*

Inpatient setting terms: **title and abstract**

Hospital* or Acute care or Secondary care or Tertiary care or Low secure or Medium secure or High secure or Secure facilit* or Forensic* or Inpatient* or Triage or (Acute hospitals or Mental health hospitals or hospitals) or Inpatients or Unit* or Ward*

Physical activity terms: **title and abstract**

Exercis* or exercise or “physical exertion” or “physical fitness” or jog* or (physical* adj3 (activ* or fit*)) or Aerobics or swim* or gym* or sport* or workout or skip* or calisthenics or e-Exercise or Bicyc* or Cycling or Parkrun or Couch to 5k or Danc*

Limits:

- Date: 2007 onwards

- English language only
