## Supplementary material for "Physical activity in adult users of inpatient mental health services: a scoping review": S2 File

**S2 File** Reference list of included reports

Research question 1: What are the correlates of physical activity in adult users of inpatient mental health services?

Ball H, Yung A, Bucci S. Staff perspectives on the barriers and facilitators to exercise implementation in inpatient mental health services: A qualitative study. Mental Health and Physical Activity. 2022 Mar 1;22:100452.

Bartlem K, Bailey J, Metse A, Asara A, Wye P, Clancy R, Wiggers J, Bowman J. Do mental health consumers want to improve their long-term disease risk behaviours? A survey of over 2000 psychiatric inpatients. International journal of mental health nursing. 2018 Jun;27(3):1032-43.

Bentzen M, Farholm A, Ivarsson A, Sørensen M. Longitudinal relations between needs satisfaction and physical activity among psychiatric patients with dual diagnoses. Advances in Mental Health. 2022 Sep 2;20(3):218-31.

Berger E, Bolstad I, Lien L, Bramness JG. The Association Between Regular Physical Activity and Depressive Symptoms Among Patients in Treatment of Alcohol and Substance Use Disorders. Substance Abuse: Research and Treatment. 2023;17.

Bergman H, Nilsson T, Andiné P, Degl’Innocenti A, Thomeé R, Gutke A. The use of physical exercise in forensic psychiatric care in Sweden: a nationwide survey. Journal of Mental Health. 2021 Jan 21:1-9.

Bierski K, Eswaran S. Finding Peace on a Psychiatric Ward with Yoga: Report on a Pilot Anthropological Study in Pondicherry, India. Ann. SBV. 2016 Jul;5(2).

Bonsaksen T. Participation in physical activity among inpatients with severe mental illness: a pilot study. International Journal of Therapy and Rehabilitation. 2011 Feb;18(2):91-9.

Brand S, Colledge F, Beeler N, Pühse U, Kalak N, Sadeghi Bahmani D, Mikoteit T, Holsboer-Trachsler E, Gerber M. The current state of physical activity and exercise programs in German-speaking, Swiss psychiatric hospitals: results from a brief online survey. Neuropsychiatric disease and treatment. 2016 Jun 3:1309-17.

Bressington D, Badnapurkar A, Inoue S, Ma HY, Chien WT, Nelson D, Gray R. Physical health care for people with severe mental illness: the attitudes, practices, and training needs of nurses in three Asian countries. International Journal of Environmental Research and Public Health. 2018 Feb;15(2):343.

Costa R, Bastos T, Probst M, Seabra A, Vilhena E, Corredeira R. Autonomous motivation and quality of life as predictors of physical activity in patients with schizophrenia. International Journal of Psychiatry in Clinical Practice. 2018 Jul 3;22(3):184-90.

Davies JL, Bagshaw R, Watt A, Hewlett P, Seage H. Staff perspectives on obesity within a Welsh secure psychiatric inpatient setting. The Journal of Mental Health Training, Education and Practice. 2023 Jan 2;18(1):44-52.

Deenik J, Kruisdijk F, Tenback D, Braakman-Jansen A, Taal E, Hopman-Rock M, Beekman A, Tak E, Hendriksen I, van Harten P. Physical activity and quality of life in long-term hospitalized patients with severe mental illness: a cross-sectional study. BMC psychiatry. 2017 Dec;17:1-1.

Deenik J, Tenback DE, Tak EC, Hendriksen IJ, van Harten PN. Improved psychosocial functioning and quality of life in inpatients with severe mental illness receiving a multidisciplinary lifestyle enhancing treatment. The MULTI study II. Mental Health and Physical Activity. 2018 Oct 1;15:145-52.

Deenik J, Tenback DE, Van Driel HF, Tak EC, Hendriksen IJ, Van Harten PN. Less medication use in inpatients with severe mental illness receiving a multidisciplinary lifestyle enhancing treatment. The MULTI Study III. Frontiers in psychiatry. 2018 Dec 18;9:707.

Deenik J, Tenback DE, Tak EC, Rutters F, Hendriksen IJ, van Harten PN. Changes in physical and psychiatric health after a multidisciplinary lifestyle enhancing treatment for inpatients with severe mental illness: The MULTI study I. Schizophrenia Research. 2019 Feb 1;204:360-7.

Deenik J, Tenback DE, Tak EC, Blanson Henkemans OA, Rosenbaum S, Hendriksen IJ, van Harten PN. Implementation barriers and facilitators of an integrated multidisciplinary lifestyle enhancing treatment for inpatients with severe mental illness: the MULTI study IV. BMC health services research. 2019 Dec;19:1-3.

Ehrbar J, Brand S, Colledge F, Donath L, Egger ST, Hatzinger M, Holsboer-Trachsler E, Imboden C, Schweinfurth N, Vetter S, Gerber M. Psychiatric in-patients are more likely to meet recommended levels of health-enhancing physical activity if they engage in exercise and sport therapy programs. Frontiers in psychiatry. 2018 Jul 20;9:322.

Emory SL, Silva SG, Christopher EJ, Edwards PB, Wahl LE. Stepping to stability and fall prevention in adult psychiatric patients. Journal of psychosocial nursing and mental health services. 2011 Dec 1;49(12):30-6.

Every-Palmer S, Huthwaite MA, Elmslie JL, Grant E, Romans SE. Long-term psychiatric inpatients perspectives on weight gain, body satisfaction, diet and physical activity: a mixed methods study. BMC psychiatry. 2018 Dec;18(1):1-9.

Fibbins H, Ward PB, Stanton R, Czsonek L, Cudmore J, Michael S, Steel Z, Rosenbaum S. Embedding an exercise professional within an inpatient mental health service: a qualitative study. Mental Health and Physical Activity. 2019 Oct 1;17:100300.

Fraser SJ, Chapman JJ, Brown WJ, Whiteford HA, Burton NW. Physical activity attitudes and preferences among inpatient adults with mental illness. International journal of mental health nursing. 2015 Oct;24(5):413-20.

Froggett L, Little R. Dance as a complex intervention in an acute mental health setting: a place ‘in-between’. British Journal of Occupational Therapy. 2012 Feb;75(2):93-9.

Furness T, Hewavasam J, Barnfield J, McKenna B, Joseph C. Adding an accredited exercise physiologist role to a new model of care at a secure extended care mental health service: a qualitative study. Journal of Mental Health. 2018 Mar 4;27(2):120-6.

Gandhi S, Gurusamy J, Damodharan D, Ganesan V. Facilitators of healthy life style behaviors in persons with schizophrenia: a qualitative feasibility pilot study. Asian Journal of Psychiatry. 2019 Feb 1;40:3-8.

Gerber M, Ehrbar J, Brand R, Antoniewicz F, Brand S, Colledge F, Donath L, Egger ST, Hatzinger M, Holsboer-Trachsler E, Imboden C. Implicit attitudes towards exercise and physical activity behaviour among in-patients with psychiatric disorders. Mental Health and Physical Activity. 2018 Oct 1;15:71-7.

Große J, Huppertz C, Röh A, Oertel V, Andresen S, Schade N, Goerke-Arndt F, Kastinger A, Schoofs N, Thomann PA, Henkel K. Step away from depression—results from a multicenter randomized clinical trial with a pedometer intervention during and after inpatient treatment of depression. European Archives of Psychiatry and Clinical Neuroscience. 2023 Aug 17:1-3.

Haddad M, Llewellyn‐Jones S, Yarnold S, Simpson A. Improving the physical health of people with severe mental illness in a low secure forensic unit: An uncontrolled evaluation study of staff training and physical health care plans. International journal of mental health nursing. 2016 Dec;25(6):554-65.

Hansen AL, Ambroziak G, Thornton D, Dahl L, Grung B. The effects of diet on levels of physical activity during winter in forensic inpatients: A randomized controlled trial. Food & Nutrition Research. 2020;64.

Hansen AL, Ambroziak G, Thornton DM, Mundt JC, Kahn RE, Dahl L, Waage L, Kattenbraker D, Grung B. Vitamin D status and physical activity during wintertime in forensic inpatients: a randomized clinical trial. Nutrients. 2021 Oct;13(10):3510.

Happell B, Scott D, Platania-Phung C, Nankivell J. Nurses' views on physical activity for people with serious mental illness. Mental Health and Physical Activity. 2012 Jun 1;5(1):4-12.

Harding SL. Direct care staff perspectives related to physical activity in mental health group homes. Journal of psychosocial nursing and mental health services. 2013 Dec 1;51(12):38-43.

Hudson NA, Mrozik JH, White R, Northend K, Moore S, Lister K, Rayner K. Community football teams for people with intellectual disabilities in secure settings: “They take you off the ward, it was like a nice day, and then you get like medals at the end”. Journal of Applied Research in Intellectual Disabilities. 2018 Mar;31(2):213-25.

Hutcheson C, Ferguson H, Nish G, Gill L. Promoting mental wellbeing through activity in a mental health hospital. British Journal of Occupational Therapy. 2010 Mar;73(3):121-8.

Kinnafick FE, Papathomas A, Regoczi D. Promoting exercise behaviour in a secure mental health setting: Healthcare assistant perspectives. International journal of mental health nursing. 2018 Dec;27(6):1776-83.

Kohn L, Christiaens W, Detraux J, De Lepeleire J, De Hert M, Gillain B, Delaunoit B, Savoye I, Mistiaen P, Jespers V. Barriers to somatic health care for persons with severe mental illness in Belgium: a qualitative study of patients' and healthcare professionals' perspectives. Frontiers in Psychiatry. 2022 Jan 26;12:798530.

Kruisdijk F, Hopman-Rock M, Beekman AT, Hendriksen IJ. Personality traits as predictors of exercise treatment adherence in major depressive disorder: lessons from a randomised clinical trial. International journal of psychiatry in clinical practice. 2020 Oct 30;24(4):380-6.

Lim KH, Morris J, Craik C. Inpatients’ perspectives of occupational therapy in acute mental health. Australian Occupational Therapy Journal. 2007 Mar;54(1):22-32.

Liu Z, Zhang Y, Sun L, Wang J, Xia L, Yang Y, Sun F, Li W, Yao X, Yang R, Liu H. Physical activity levels associated with insomnia and depressive symptoms in middle-aged and elderly patients with chronic schizophrenia. Frontiers in Psychiatry. 2023 Jan 6;13:1045398.

Martland R, Gaughran F, Stubbs B, Onwumere J. Perspectives on implementing HIIT interventions for service users in inpatient mental health settings: a qualitative study investigating patient, carer and staff attitudes. Journal of Affective Disorders. 2021 Mar 15;283:198-206.

Mateo-Urdiales A, Michael M, Simpson C, Beenstock J. Evaluation of a participatory approach to improve healthy eating and physical activity in a secure mental health setting. Journal of Public Mental Health. 2020 Apr 7;19(4):301-9.

Moss K, Meurk C, Steele ML, Heffernan E. Physical Health and Activity of Inpatients under Forensic Mental Health Care: A Cross-Sectional Survey and Audit of Patients in a High Secure Setting in Queensland, Australia. International Journal of Forensic Mental Health. 2022 Dec 3:1-2.

Mugisha J, De Hert M, Knizek BL, Kwiringira J, Kinyanda E, Byansi W, van Winkel R, Myin-Germeys I, Stubbs B, Vancampfort D. Health care professionals perspectives on physical activity within the Ugandan mental health care system. Mental Health and Physical Activity. 2019 Mar 1;16:1-7.

Perham A, Accordino M. Exercise and functioning level of individuals with severe mental illness: a comparison of two groups. Journal of Mental Health Counseling. 2007 Oct 1;29(4):350-62.

Prebble K, Kidd J, O'Brien A, Carlyle D, McKenna B, Crowe M, Deering D, Gooder C. Implementing and maintaining nurse-led healthy living programs in forensic inpatient settings: an illustrative case study. Journal of the American Psychiatric Nurses Association. 2011 Mar;17(2):127-38.

Reynolds P, Field L. Audit of group-based activities in an inpatient assessment and treatment unit for individuals with learning disabilities. British Journal of Learning Disabilities. 2013 Dec;41(4):273-9.

Rezaie L, Shafaroodi N, Philips D. The barriers to participation in leisure time physical activities among Iranian women with severe mental illness: A qualitative study. Mental Health and Physical Activity. 2017 Oct 1;13:171-7.

Robson D, Haddad M, Gray R, Gournay K. Mental health nursing and physical health care: A cross-sectional study of nurses' attitudes, practice, and perceived training needs for the physical health care of people with severe mental illness. International Journal of Mental Health Nursing. 2013 Oct;22(5):409-17.

Roempler J, Petzold MB, Bendau A, Plag J, Ströhle A. Tracking changes in physical activity during inpatient treatment in a psychiatric clinic in Germany by asking two simple questions. European Archives of Psychiatry and Clinical Neuroscience. 2023 Feb 11:1-2.

Rogers E, Papathomas A, Kinnafick FE. Inpatient perspectives on physical activity in a secure mental health setting. Psychology of Sport and Exercise. 2021 Jan 1;52:101827.

Seet V, Abdin E, Asharani PV, Lee YY, Roystonn K, Wang P, Devi F, Cetty L, Teh WL, Verma S, Mok YM. Physical activity, sedentary behaviour and smoking status among psychiatric patients in Singapore: a cross-sectional study. BMC psychiatry. 2021 Dec;21(1):1-0.

Shin S, Yeom CW, Shin C, Shin JH, Jeong JH, Shin JU, Lee YR. Activity monitoring using a mHealth device and correlations with psychopathology in patients with chronic schizophrenia. Psychiatry Research. 2016 Dec 30;246:712-8.

Sørensen M, Bentzen M, Farholm A. Motivational physical activity intervention for psychiatric inpatients: A two phased single-cases experimental study.

Sørensen M, Bentzen M, Farholm A. Lessons learned from a physical activity intervention in psychiatric treatment: Patient, staff, and leader perspectives. Frontiers in psychiatry. 2020 Mar 2;11:87.

Soundy A, Stubbs B, Probst M, Hemmings L, Vancampfort D. Barriers to and facilitators of physical activity among persons with schizophrenia: a survey of physical therapists. Psychiatric Services. 2014 May;65(5):693-6.

Soundy A, Freeman P, Stubbs B, Probst M, Vancampfort D. The value of social support to encourage people with schizophrenia to engage in physical activity: an international insight from specialist mental health physiotherapists. Journal of Mental Health. 2014 Oct 1;23(5):256-60.

Stanton R, Reaburn P, Happell B. Barriers to exercise prescription and participation in people with mental illness: the perspectives of nurses working in mental health. Journal of Psychiatric and Mental Health Nursing. 2015 Aug;22(6):440-8.

Stanton R, Happell B, Reaburn P. Investigating the exercise prescription practices of nurses working in inpatient mental health settings. International Journal of Mental Health Nursing. 2015 Apr;24(2):112-20.

Stubbs B, Soundy A, Probst M, De Hert M, De Herdt A, Parker A, Vancampfort D. The assessment, benefits and delivery of physical activity in people with schizophrenia: a survey of members of the International Organization of Physical Therapists in Mental Health. Physiotherapy Research International. 2014 Dec;19(4):248-56.

Tetlie T, Heimsnes MC, Almvik R. Using exercise to treat patients with severe mental illness: how and why? J Psychosoc Nurs Ment Health Serv. 2009 Feb;47(2):32-40. doi: 10.3928/02793695-20090201-14.

Ussher M, Stanbury L, Cheeseman V, Faulkner G. Physical activity preferences and perceived barriers to activity among persons with severe mental illness in the United Kingdom. Psychiatr Serv. 2007 Mar;58(3):405-8. doi: 10.1176/ps.2007.58.3.405.

Vancampfort D, De Hert M, Sweers K, De Herdt A, Detraux J, Probst M. Diabetes, physical activity participation and exercise capacity in patients with schizophrenia. Psychiatry and Clinical Neurosciences. 2013 Sep;67(6):451-6.

Vancampfort D, Stubbs B, Venigalla SK, Probst M. Adopting and maintaining physical activity behaviours in people with severe mental illness: The importance of autonomous motivation. Preventive medicine. 2015 Dec 1;81:216-20.

Vancampfort D, Madou T, Moens H, De Backer T, Vanhalst P, Helon C, Naert P, Rosenbaum S, Stubbs B, Probst M. Could autonomous motivation hold the key to successfully implementing lifestyle changes in affective disorders? A multicentre cross sectional study. Psychiatry Res. 2015 Jul 30;228(1):100-6. doi: 10.1016/j.psychres.2015.04.021.

Vancampfort D, De Hert M, Stubbs B, Ward PB, Rosenbaum S, Soundy A, Probst M. Negative symptoms are associated with lower autonomous motivation towards physical activity in people with schizophrenia. Comprehensive Psychiatry. 2015 Jan 1;56:128-32.

Vancampfort D, Sienaert P, Wyckaert S, De Hert M, Stubbs B, Probst M. Sitting time, physical fitness impairments and metabolic abnormalities in people with bipolar disorder: An exploratory study. Psychiatry Res. 2016 Aug 30;242:7-12. doi: 10.1016/j.psychres.2016.05.023.

Vancampfort D, Van Damme T, Probst M, Vandael H, Hallgren M, Mutamba BB, Nabanoba J, Basangwa D, Mugisha J. Motives for physical activity in the adoption and maintenance of physical activity in men with alcohol use disorders. Psychiatry Research. 2018 Mar 1;261:522-6.

Walker K, Yates J, Dening T, Völlm B, Tomlin J, Griffiths C. Staff perspectives on barriers to and facilitators of quality of life, health, wellbeing, recovery and reduced risk for older forensic mental-health patients: A qualitative interview study. Journal of Health Services Research & Policy. 2022 Oct;27(4):287-300.

Wright A, Cattan M. Physical activity and the management of depression. Working with Older People. 2009 Apr 13;13(1):15-8.

Wynaden D, Barr L, Omari O, Fulton A. Evaluation of service users' experiences of participating in an exercise programme at the Western Australian State Forensic Mental Health Services. International journal of mental health nursing. 2012 Jun;21(3):229-35.

Research question 2: What are the characteristics and findings of studies of physical activity interventions for this population?

Abrantes AM, Meshesha LZ, E Blevins C, Battle CL, Lindsay C, Marsh E, Feltus S, Buman M, Agu E, Stein M. A Smartphone Physical Activity App for Patients in Alcohol Treatment: Single-Arm Feasibility Trial. JMIR Formative Research. 2022 Oct 19;6(10):e35926.

Areshtanab HN, Ebrahimi H, Abdi M, Mohammadian R, Asl AM, Piri S. The Effect of Aerobic Exercise on the Quality of Life of Male Patients Who Suffer from Chronic Schizophrenia: Double-Blind, Randomized Control Trial. Iranian Journal of Psychiatry and Behavioral Sciences. 2020 Dec 31;14(4).

Babson KA, Heinz AJ, Ramirez G, Puckett M, Irons JG, Bonn-Miller MO, Woodward SH. The interactive role of exercise and sleep on veteran recovery from symptoms of PTSD. Mental Health and Physical Activity. 2015 Mar 1;8:15-20.

Bacon N, Farnworth L, Boyd R. The use of the Wii Fit in forensic mental health: exercise for people at risk of obesity. British Journal of Occupational Therapy. 2012 Feb;75(2):61-8.

Bentzen M, Farholm A, Ivarsson A, Sørensen M. Longitudinal relations between needs satisfaction and physical activity among psychiatric patients with dual diagnoses. Advances in Mental Health. 2022 Sep 2;20(3):218-31.

Bernard PP, Esseul EC, Raymond L, Dandonneau L, Xambo JJ, Carayol MS, Ninot GJ. Counseling and exercise intervention for smoking reduction in patients with schizophrenia: a feasibility study. Archives of psychiatric nursing. 2013 Feb 1;27(1):23-31.

Bierski K, Eswaran S. Finding Peace on a Psychiatric Ward with Yoga: Report on a Pilot Anthropological Study in Pondicherry, India. Ann. SBV. 2016 Jul;5(2).

Biondo J, Gerber N. Single-session dance/movement therapy for people with acute schizophrenia: Development of a treatment protocol. American Journal of Dance Therapy. 2020 Dec;42(2):277-95.

Biondo J, Gerber N, Bradt J, Du W, Goodill S. Single-session dance/movement therapy for thought and behavioral dysfunction associated with schizophrenia: A mixed methods feasibility study. The Journal of Nervous and Mental Disease. 2021 Feb 1;209(2):114-22.

Bjerg Christensen J, Lassen IS, Helles Carlsen A, Strazek S, Nyboe L. Physical therapy for reducing arousal and mechanical restraint among in-patients with mania. Nordic Journal of Psychiatry. 2021 Jan 2;75(1):49-53.

Bonsaksen T. Participation in physical activity among inpatients with severe mental illness: a pilot study. International Journal of Therapy and Rehabilitation. 2011 Feb;18(2):91-9.

Borge L, Røssberg JI, Sverdrup S. Cognitive milieu therapy and physical activity: experiences of mastery and learning among patients with dual diagnosis. Journal of psychiatric and mental health nursing. 2013 Dec;20(10):932-42.

Brand S, Colledge F, Ludyga S, Emmenegger R, Kalak N, Sadeghi Bahmani D, Holsboer-Trachsler E, Pühse U, Gerber M. Acute bouts of exercising improved mood, rumination and social interaction in inpatients with mental disorders. Frontiers in psychology. 2018 Mar 13;9:249.

Bueno-Antequera J, Oviedo-Caro MÁ, Munguía-Izquierdo D. Feasibility and effects of an exercise-based intervention in prison inmates with psychiatric disorders: the PsychiActive project randomized controlled trial. Clinical Rehabilitation. 2019 Oct;33(10):1661-71.

Bukar NK, Eberhardt LM, Davidson J. East meets west in psychiatry: Yoga as an adjunct therapy for management of anxiety. Archives of psychiatric nursing. 2019 Aug 1;33(4):371-6.

Bürge E, Berchtold A, Maupetit C, Bourquin NM, Von Gunten A, Ducraux D, Zumbach S, Peeters A, Kuhne N. Does physical exercise improve ADL capacities in people over 65 years with moderate or severe dementia hospitalized in an acute psychiatric setting? A multisite randomized clinical trial. International psychogeriatrics. 2017 Feb;29(2):323-32.

Buschert V, Prochazka D, Bartl H, Diemer J, Malchow B, Zwanzger P, Brunnauer A. Effects of physical activity on cognitive performance: a controlled clinical study in depressive patients. European archives of psychiatry and clinical neuroscience. 2019 Aug 1;269:555-63.

Chaves LG, Gama DR, Castro JB, Oliveira KR, Vale RG. Cortisol and serotonin levels in schizophrenic inpatients undergoing aerobic training. Revista Brasileira de Medicina do Esporte. 2020 Jul 29;26:307-11.

Cisse A, Giles C, Salloum IM. Yoga in the Yard. Psychiatric Services. 2017 Sep 1;68(9):980-.

Cody R, Christensen M, Kreppke JN, Faude O, Gerber M, Nicca D. The experience of a physical activity counseling intervention among people with major depression within the PACINPAT trial: A reflexive thematic analysis. Mental Health and Physical Activity. 2022 Oct 1;23:100464.

Cody R, Beck J, Brand S, Donath L, Faude O, Hatzinger M, Imboden C, Kreppke JN, Lang UE, Ludyga S, Mans S. Short-term outcomes of physical activity counseling in in-patients with Major Depressive Disorder: Results from the PACINPAT randomized controlled trial. Frontiers in Psychiatry. 2023;13:3074.

Cordes J, Thünker J, Regenbrecht G, Zielasek J, Correll CU, Schmidt-Kraepelin C, Lange-Asschenfeldt C, Agelink MW, Kahl KG, Gaebel W, Klimke A. Can an early weight management program (WMP) prevent olanzapine (OLZ)-induced disturbances in body weight, blood glucose and lipid metabolism? Twenty-four-and 48-week results from a 6-month randomized trial. The World Journal of Biological Psychiatry. 2014 Apr 1;15(3):229-41.

Cormac I, Hallford S, Hart L, Creasey S, Ferriter M. Evaluation of an integrated weight management and fitness programme in a high-security psychiatric setting. Psychiatric Bulletin. 2008 Mar;32(3):95-8.

Cormac I, Ferriter M, Buchan S. Follow-up study of an integrated weight management and fitness programme. Mental Health Review Journal. 2013 Mar 22;18(1):14-20.

Curcic D, Stojmenovic T, Djukic-Dejanovic S, Dikic N, Vesic-Vukasinovic M, Radivojevic N, Andjelkovic M, Borovcanin M, Djokic G. Positive impact of prescribed physical activity on symptoms of schizophrenia: randomized clinical trial. Psychiatria Danubina. 2017 Dec 4;29(4):459-65.

Dai Y, Ding H, Lu X, Wu X, Xu C, Jiang T, Ming L, Xia Z, Song C, Shen H, Hao W. CCRT and aerobic exercise: a randomised controlled study of processing speed, cognitive flexibility, and serum BDNF expression in schizophrenia. Schizophrenia. 2022 Oct 20;8(1):84.

Deenik J, Tenback DE, Tak EC, Hendriksen IJ, van Harten PN. Improved psychosocial functioning and quality of life in inpatients with severe mental illness receiving a multidisciplinary lifestyle enhancing treatment. The MULTI study II. Mental Health and Physical Activity. 2018 Oct 1;15:145-52.

Deenik J, Tenback DE, Van Driel HF, Tak EC, Hendriksen IJ, Van Harten PN. Less medication use in inpatients with severe mental illness receiving a multidisciplinary lifestyle enhancing treatment. The MULTI Study III. Frontiers in psychiatry. 2018 Dec 18;9:707.

Deenik J, Tenback DE, Tak EC, Blanson Henkemans OA, Rosenbaum S, Hendriksen IJ, van Harten PN. Implementation barriers and facilitators of an integrated multidisciplinary lifestyle enhancing treatment for inpatients with severe mental illness: the MULTI study IV. BMC health services research. 2019 Dec;19:1-3.

Deenik J, Tenback DE, Tak EC, Rutters F, Hendriksen IJ, van Harten PN. Changes in physical and psychiatric health after a multidisciplinary lifestyle enhancing treatment for inpatients with severe mental illness: The MULTI study I. Schizophrenia Research. 2019 Feb 1;204:360-7.

Dodd KJ, Duffy S, Stewart JA, Impey J, Taylor N. A small group aerobic exercise programme that reduces body weight is feasible in adults with severe chronic schizophrenia: a pilot study. Disability and Rehabilitation. 2011 Jan 1;33(13-14):1222-9.

Dürmüs P, Vardar ME, Kaya O, Tayfur P, Süt N, Vardar SA. Evaluation of the Effects of High Intensity Interval Training on Cytokine Levels and Clinical Course in Treatment of Opioid Use Disorder. Turk Psikiyatri Dergisi. 2020 Sep 1;31(3).

Ellingsen MM, Clausen T, Johannesen SL, Martinsen EW, Hallgren M. Effects of acute exercise on affect, anxiety, and self-esteem in poly-substance dependent inpatients. European addiction research. 2023 Aug 21;29(4):285-93.

Elmasry NM, Ali EF, Abdelmoez RM. Effect of lifestyle modification on schizophrenia patients on antipsychotics. Egyptian Journal of Psychiatry. 2021 May 1;42(2):100.

Emory SL, Silva SG, Christopher EJ, Edwards PB, Wahl LE. Stepping to stability and fall prevention in adult psychiatric patients. Journal of psychosocial nursing and mental health services. 2011 Dec 1;49(12):30-6.

Estoque HV. Effects of Steps to Success Exercise Program in Weight Management For Psychiatric Patients Taking Atypical Antipsychotic Drugs. Journal Of Nursing Practice. 2020 Oct 10;4(1):84-96.

Firth J, Carney R, Pownall M, French P, Elliott R, Cotter J, Yung AR. Challenges in implementing an exercise intervention within residential psychiatric care: A mixed methods study. Mental health and physical activity. 2017 Mar 1;12:141-6.

Flemmen G, Unhjem R, Wang E. High-intensity interval training in patients with substance use disorder. BioMed research international. 2014 Oct;2014.

Froggett L, Little R. Dance as a complex intervention in an acute mental health setting: a place “in-between”. British Journal of Occupational Therapy. 2012 Feb;75(2):93-9.

Fruehauf A, Niedermeier M, Elliott LR, Ledochowski L, Marksteiner J, Kopp M. Acute effects of outdoor physical activity on affect and psychological well-being in depressed patients: A preliminary study. Mental Health and Physical Activity. 2016 Mar 1;10:4-9.

Furzer BJ, Wright KE, Edoo A, Maiorana A. Move your mind: embedding accredited exercise physiology services within a hospital-based mental health service. Australasian Psychiatry. 2021 Feb;29(1):52-6.

Gerber M, Minghetti A, Beck J, Zahner L, Donath L. Sprint interval training and continuous aerobic exercise training have similar effects on exercise motivation and affective responses to exercise in patients with major depressive disorders: a randomized controlled trial. Frontiers in psychiatry. 2018 Dec 21;9:694.

Gholipour A, Abolghasemi SH, Gholinia K, Taheri S. Token reinforcement therapeutic approach is more effective than exercise for controlling negative symptoms of schizophrenic patients: a randomized controlled trial. International Journal of Preventive Medicine. 2012 Jul;3(7):466.

Göhner W, Dietsche C, Fuchs R. Increasing physical activity in patients with mental illness—a randomized controlled trial. Patient education and counseling. 2015 Nov 1;98(11):1385-92.

Gorain RK, Ramu R, Sinha P, Govindan R. Impact of structured physical activity program on the level of functional ability of Persons with Mental Illness. Journal of Education and Health Promotion. 2022 Jan 1;11(1):226.

Guan H, Zhou Z, Li X, Pan Y, Zou Z, Meng X, Guan K, Zhang L, Li Z, Li X, Wei B. Dance/movement therapy for improving balance ability and bone mineral density in long-term patients with schizophrenia: a randomized controlled trial. Schizophrenia. 2023 Jul 31;9(1):47.

Hanssen H, Minghetti A, Faude O, Schmidt-Trucksäss A, Zahner L, Beck J, Donath L. Effects of endurance exercise modalities on arterial stiffness in patients suffering from unipolar depression: a randomized controlled trial. Frontiers in psychiatry. 2018 Jan 22;8:311.

Hanssen H, Minghetti A, Faude O, Schmidt-Trucksäss A, Zahner L, Beck J, Donath L. Effects of different endurance exercise modalities on retinal vessel diameters in unipolar depression. Microvascular research. 2018 Nov 1;120:111-6.

Haussleiter IS, Bolsinger B, Assion HJ, Juckel G. Adjuvant guided exercise therapy versus self-organized activity in patients with major depression. The Journal of Nervous and Mental Disease. 2020 Dec 1;208(12):982-8.

Heggelund J, Nilsberg GE, Hoff J, Morken G, Helgerud J. Effects of high aerobic intensity training in patients with schizophrenia: a controlled trial. Nordic journal of psychiatry. 2011 Sep 1;65(4):269-75.

Heissel A, Vesterling A, White SA, Kallies G, Behr D, Arafat AM, Reischies FM, Heinzel S, Budde H. Feasibility of an exercise program for older depressive inpatients. GeroPsych. 2015 Nov 26.

Hjorth P, Davidsen AS, Kilian R, Pilgaard Eriksen S, Jensen SO, Sørensen HØ, Munk-Jørgensen P. Improving the physical health of long-term psychiatric inpatients. Australian & New Zealand Journal of Psychiatry. 2014 Sep;48(9):861-70.

Ho CW, Chan SC, Wong JS, Cheung WT, Chung DW, Lau TF. Effect of aerobic exercise training on Chinese population with mild to moderate depression in Hong Kong. Rehabilitation Research and Practice. 2014 Oct;2014.

Ho RT, Fong TC, Wan AH, Au-Yeung FS, Wong CP, Ng WY, Cheung IK, Lo PH, Ng SM, Chan CL, Chen EY. A randomized controlled trial on the psychophysiological effects of physical exercise and Tai-chi in patients with chronic schizophrenia. Schizophrenia research. 2016 Mar 1;171(1-3):42-9.

Hudson NA, Mrozik JH, White R, Northend K, Moore S, Lister K, Rayner K. Community football teams for people with intellectual disabilities in secure settings: “They take you off the ward, it was like a nice day, and then you get like medals at the end”. Journal of Applied Research in Intellectual Disabilities. 2018 Mar;31(2):213-25.

Hutcheson C, Ferguson H, Nish G, Gill L. Promoting mental wellbeing through activity in a mental health hospital. British Journal of Occupational Therapy. 2010 Mar;73(3):121-8.

Hutchison SL, Terhorst L, Murtaugh S, Gross S, Kogan JN, Shaffer SL. Effectiveness of a staff promoted wellness program to improve health in residents of a mental health long-term care facility. Issues in mental health nursing. 2016 Apr 2;37(4):257-64.

Ikai S, Uchida H, Mizuno Y, Tani H, Nagaoka M, Tsunoda K, Mimura M, Suzuki T. Effects of chair yoga therapy on physical fitness in patients with psychiatric disorders: A 12-week single-blind randomized controlled trial. Journal of Psychiatric Research. 2017 Nov 1;94:194-201.

Imboden C, Gerber M, Beck J, Holsboer-Trachsler E, Pühse U, Hatzinger M. Aerobic exercise or stretching as add-on to inpatient treatment of depression: Similar antidepressant effects on depressive symptoms and larger effects on working memory for aerobic exercise alone. Journal of affective disorders. 2020 Nov 1;276:866-76.

Jacquart SD, Marshak HH, Dos Santos H, Luu SM, Berk LS, McMahon PT, Riggs M. The effects of simultaneous exercise and psychotherapy on depressive symptoms in inpatient, psychiatric older adults. Advances in mind-body medicine. 2014 Sep 1;28(4):8-17.

Jasińska-Mikołajczyk A, Drews K, Domaszewska K, Kolasa G, Konofalska M, Jowik K, Skibińska M, Rybakowski F. The effect of physical activity on neurotrophin concentrations and cognitive control in patients with a depressive episode. Frontiers in Psychiatry. 2022 Apr 25;13:777394.

Jo G, Rossow-Kimball B, Park G, Lee Y. Effects of virtual reality exercise for Korean adults with schizophrenia in a closed ward. Journal of exercise rehabilitation. 2018 Feb;14(1):39.

Kahl KG, Kerling A, Tegtbur U, Gützlaff E, Herrmann J, Borchert L, Ates Z, Westhoff-Bleck M, Hueper K, Hartung D. Effects of additional exercise training on epicardial, intra-abdominal and subcutaneous adipose tissue in major depressive disorder: A randomized pilot study. Journal of affective disorders. 2016 Mar 1;192:91-7.

Kampragkou C, Iakovidis P, Kampragkou E, Kellis E. Effects of a 12-week aerobic exercise program combined with music therapy and memory exercises on cognitive and functional ability in people with middle type of Alzheimer's disease. International Journal of Physiotherapy. 2017 Oct 1;4(5):262-8.

Kerling A, Tegtbur U, Gützlaff E, Kück M, Borchert L, Ates Z, Von Bohlen A, Frieling H, Hüper K, Hartung D, Schweiger U. Effects of adjunctive exercise on physiological and psychological parameters in depression: a randomized pilot trial. Journal of affective disorders. 2015 May 15;177:1-6.

Kerling A, von Bohlen A, Kück M, Tegtbur U, Grams L, Haufe S, Gützlaff E, Kahl KG. Exercise therapy improves aerobic capacity of inpatients with major depressive disorder. Brain and behavior. 2016 Jun;6(6):e00469.

Kerling A, Kück M, Tegtbur U, Grams L, Weber-Spickschen S, Hanke A, Stubbs B, Kahl KG. Exercise increases serum brain-derived neurotrophic factor in patients with major depressive disorder. Journal of affective disorders. 2017 Jun 1;215:152-5.

Kerling A, Hartung D, Stubbs B, Kück M, Tegtbur U, Grams L, Weber-Spickschen TS, Kahl KG. Impact of aerobic exercise on muscle mass in patients with major depressive disorder: a randomized controlled trial. Neuropsychiatric disease and treatment. 2018 Aug 6:1969-74.

Kim HJ, Song BK, So B, Lee O, Song W, Kim Y. Increase of circulating BDNF levels and its relation to improvement of physical fitness following 12 weeks of combined exercise in chronic patients with schizophrenia: a pilot study. Psychiatry research. 2014 Dec 30;220(3):792-6.

Kim YS, Song BK, Oh JS, Woo SS. Aerobic exercise improves gastrointestinal motility in psychiatric inpatients. World Journal of Gastroenterology: WJG. 2014 Aug 8;20(30):10577.

Kleinstäuber M, Reuter M, Doll N, Fallgatter AJ. Rock climbing and acute emotion regulation in patients with major depressive disorder in the context of a psychological inpatient treatment: a controlled pilot trial. Psychology research and behavior management. 2017 Aug 16:277-81.

Knubben K, Reischies FM, Adli M, Schlattmann P, Bauer M, Dimeo F. A randomised, controlled study on the effects of a short-term endurance training programme in patients with major depression. British journal of sports medicine. 2007 Jan 1;41(1):29-33.

Koch SC, Morlinghaus K, Fuchs T. The joy dance: Specific effects of a single dance intervention on psychiatric patients with depression. The arts in Psychotherapy. 2007 Jan 1;34(4):340-9.

Koch SC, Wirtz G, Harter C, Weisbrod M, Winkler F, Pröger A, Herpertz SC. Embodied self in trauma and self-harm: a pilot study of effects of flamenco therapy on traumatized inpatients. Journal of loss and trauma. 2019 Aug 18;24(5-6):441-59.

Korman, N.H., Shah, S., Suetani, S., Kendall, K., Rosenbaum, S., Dark, F., Nadareishvili, K. and Siskind, D., 2018. Evaluating the feasibility of a pilot exercise intervention implemented within a residential rehabilitation unit for people with severe mental illness: GO HEART:(Group Occupational Health Exercise and Rehabilitation Treatment). Frontiers in Psychiatry, 9, p.343.

Korman N, Fox H, Skinner T, Dodd C, Suetani S, Chapman J, Parker S, Dark F, Collins C, Rosenbaum S, Siskind D. Feasibility and acceptability of a student-led lifestyle (diet and exercise) intervention within a residential rehabilitation setting for people with severe mental illness, GO HEART (Group Occupation, Health, Exercise And Rehabilitation Treatment). Frontiers in Psychiatry. 2020 Apr 28;11:319.

Kurebayashi Y, Mori K, Otaki J. Effects of mild-intensity physical exercise on neurocognition in inpatients with schizophrenia: A pilot randomized controlled trial. Perspectives in Psychiatric Care. 2022 Jul;58(3):1037-47.

Lan YL, Ping LY, Su LW, Chen CC. The Impact of Health Promotion Activities on the Physiological, Psychological, and Social Functions of Inpatients With Chronic Mental Illness. Psychiatry Investigation. 2022 Mar;19(3):171.

Landin M, Palmer C, Paul N, Shahrjerdi P. Improving physical health monitoring and interventions in a learning disabilities forensic psychiatric secure service. Int J Risk Saf Med. 2022;33(S1):S85-S90. doi: 10.3233/JRS-227030.

Lee HL, Hwang EJ, Wu SL, Hsu WC. Appraising Psychiatric Care From a Different Angle: Occupational Therapy Activities and Cardiorespiratory Fitness for Inpatients With Chronic Mental Illness. The American Journal of Occupational Therapy. 2022 Jul 1;76(4).

Legrand FD, Neff EM. Efficacy of exercise as an adjunct treatment for clinically depressed inpatients during the initial stages of antidepressant pharmacotherapy: an open randomized controlled trial. Journal of affective disorders. 2016 Feb 1;191:139-44.

Legrand FD, Lallement D, Kasmi S. Physical activity can reduce hopelessness among women admitted to psychiatric short stay unit following a suicide crisis. Journal of psychiatric research. 2022 Nov 1;155:567-71.

Li M, Fang J, Gao Y, Wu Y, Shen L, Yusubujiang Y, Luo J. Baduanjin mind-body exercise improves logical memory in long-term hospitalized patients with schizophrenia: A randomized controlled trial. Asian Journal of Psychiatry. 2020 Jun 1;51:102046.

Lim KH, Morris J, Craik C. Inpatients’ perspectives of occupational therapy in acute mental health. Australian Occupational Therapy Journal. 2007 Mar;54(1):22-32.

Lindenmayer JP, Khan A, Wance D, Maccabee N, Kaushik S, Kaushik S. Outcome evaluation of a structured educational wellness program in patients with severe mental illness. The Journal of clinical psychiatry. 2009 Sep 22;70(10):13562.

Lippi S, Petit L. Subverting space: An exploration of a dance therapy workshop apparatus for schizophrenics. The Psychoanalytic Review. 2017 Apr;104(2):231-52.

Liu B. MUSIC AND DANCE ON THE TREATMENT OF DEPRESSIVE PSYCHOSIS. Psychiatria Danubina. 2022 Apr 29;34(suppl 1):908-13.

Loh SY, Abdullah A, Bakar AK, Thambu M, Jaafar NR. Structured walking and chronic institutionalized schizophrenia inmates: a pilot RCT study on quality of life. Global journal of health science. 2016 Jan;8(1):238.

Long C, Mason F. Improving health and wellbeing in women's secure services: Physical activity, appearance, self-care and body image. Ethnicity and Inequalities in Health and Social Care. 2014 Dec 9;7(4):178-86.

Long C, West R, Rigg S, Spickett R, Murray L, Savage P, Butler S, Stillman SK, Dolley O. Increasing physical activity in a secure psychiatric service for women. Mental Health Review Journal. 2015 Sep 14;20(3):144-55.

Magni LR, Ferrari C, Rossi G, Staffieri E, Uberti A, Lamonaca D, Boggian I, Merlin S, Primerano G, Mombrini A, Poli R. Superwellness Program: a cognitive-behavioral therapy-based group intervention to reduce weight gain in patients treated with antipsychotic drugs. Brazilian Journal of Psychiatry. 2017 Mar 13;39:244-51.

Malchow B, Keller K, Hasan A, Dörfler S, Schneider-Axmann T, Hillmer-Vogel U, Honer WG, Schulze TG, Niklas A, Wobrock T, Schmitt A. Effects of endurance training combined with cognitive remediation on everyday functioning, symptoms, and cognition in multiepisode schizophrenia patients. Schizophrenia Bulletin. 2015 Jul 1;41(4):847-58.

Malchow B, Keeser D, Keller K, Hasan A, Rauchmann BS, Kimura H, Schneider-Axmann T, Dechent P, Gruber O, Ertl-Wagner B, Honer WG. Effects of endurance training on brain structures in chronic schizophrenia patients and healthy controls. Schizophrenia Research. 2016 Jun 1;173(3):182-91.

Manjunath RB, Varambally S, Thirthalli J, Basavaraddi IV, Gangadhar BN. Efficacy of yoga as an add-on treatment for in-patients with functional psychotic disorder. Indian journal of psychiatry. 2013 Jul 1;55(Suppl 3):S374-8.

Mateo-Urdiales A, Michael M, Simpson C, Beenstock J. Evaluation of a participatory approach to improve healthy eating and physical activity in a secure mental health setting. Journal of Public Mental Health. 2020 Apr 7;19(4):301-9.

Mazyarkin Z, Peleg T, Golani I, Sharony L, Kremer I, Shamir A. Health benefits of a physical exercise program for inpatients with mental health; a pilot study. Journal of psychiatric research. 2019 Jun 1;113:10-6.

McCartney D, Isik AD, Rooney K, Arnold JC, Bartlett DJ, Murnion B, Richards E, Arkell TR, Lintzeris N, McGregor IS. The effect of daily aerobic cycling exercise on sleep quality during inpatient cannabis withdrawal: A randomised controlled trial. Journal of Sleep Research. 2021 Jun;30(3):e13211.

Melamed Y, Stein-Reisner O, Gelkopf M, Levi G, Sivan T, Ilievici G, Rosenberg R, Weizman A, Bleich A. Multi-modal weight control intervention for people with persistent mental disorders. Psychiatric Rehabilitation Journal. 2008;31(3):194.

Methapatara W, Srisurapanont M. Pedometer walking plus motivational interviewing program for Thai schizophrenic patients with obesity or overweight: A 12-week, randomized, controlled trial. Psychiatry and clinical neurosciences. 2011 Jun;65(4):374-80.

Minghetti A, Faude O, Hanssen H, Zahner L, Gerber M, Donath L. Sprint interval training (SIT) substantially reduces depressive symptoms in major depressive disorder (MDD): a randomized controlled trial. Psychiatry research. 2018 Jul 1;265:292-7.

Ng F, Dodd S, Jacka FN, Leslie E, Berk M. Effects of a walking program in the psychiatric in-patient treatment setting: a cohort study. Health Promotion Journal of Australia. 2007;18(1):39-42.

Ng F, Dodd S, Berk M. The effects of physical activity in the acute treatment of bipolar disorder: a pilot study. Journal of affective disorders. 2007 Aug 1;101(1-3):259-62.

Niedermeier M, Ledochowski L, Leitner H, Zingerle H, Kopp M. Acute effects of a single bout of walking on affective responses in patients with major depressive disorder. International journal of environmental research and public health. 2021 Feb;18(4):1524.

Oertel-Knöchel V, Mehler P, Thiel C, Steinbrecher K, Malchow B, Tesky V, Ademmer K, Prvulovic D, Banzer W, Zopf Y, Schmitt A. Effects of aerobic exercise on cognitive performance and individual psychopathology in depressive and schizophrenia patients. European archives of psychiatry and clinical neuroscience. 2014 Oct;264:589-604.

Panagiotounis F, Hassandra M, Krommidas C, Theodorakis Y. Effects of an exercise theory-based intervention program on craving during the early stage of adults' SUD treatment. Mental Health and Physical Activity. 2022 Oct 1;23:100463.

Perham A, Accordino M. Exercise and functioning level of individuals with severe mental illness: a comparison of two groups. Journal of Mental Health Counseling. 2007 Oct 1;29(4):350-62.

Pitkänen A, Alanen HM, Kampman O, Suontaka-Jamalainen K, Leinonen E. Implementing physical exercise and music interventions for patients suffering from dementia on an acute psychogeriatric inpatient ward. Nordic journal of psychiatry. 2019 Oct 3;73(7):401-8.

Polanco-Zuleta KM, Medina-Corrales M, Mendoza-Farías FJ, Lozano CC, Tristán J, Pappous AS, López-Walle JM. Effects of a dance program on psychophysiological variables in hospitalized patients with depression: A mixed model approach. The Arts in Psychotherapy. 2021 Nov 1;76:101857.

Prebble K, Kidd J, O'Brien A, Carlyle D, McKenna B, Crowe M, Deering D, Gooder C. Implementing and maintaining nurse-led healthy living programs in forensic inpatient settings: an illustrative case study. Journal of the American Psychiatric Nurses Association. 2011 Mar;17(2):127-38.

Rahmani J, Khosravani F. Effects of Group Exercise (Sports Team) on Hospitalized Depressed Patients. Procedia-Social and Behavioral Sciences. 2015 May 13;185:104-8.

Rauchmann BS, Ghaseminejad F, Keeser D, Keller-Varady K, Schneider-Axmann T, Takahashi S, Karali T, Helms G, Dechent P, Maurus I, Hasan A. The impact of endurance training and table soccer on brain metabolites in schizophrenia. Brain Imaging and Behavior. 2020 Apr;14:515-26.

Rebar AL, Faulkner G, Stanton R. An exploratory study examining the core affect hypothesis of the anti-depressive and anxiolytic effects of physical activity. Mental Health and Physical Activity. 2015 Oct 1;9:55-8.

Reimer V, Kanning MK. Does sports therapy affect momentary affective states? Feasibility of intensive longitudinal case studies in forensic psychiatry. Frontiers in Psychiatry. 2023 May 12;14:847.

Reynolds P, Field L. Audit of group-based activities in an inpatient assessment and treatment unit for individuals with learning disabilities. British Journal of Learning Disabilities. 2013 Dec;41(4):273-9.

Ringen PA, Falk RS, Antonsen B, Faerden A, Mamen A, Rognli EB, Solberg DK, Martinsen EW, Andreassen OA. Using motivational techniques to reduce cardiometabolic risk factors in long term psychiatric inpatients: a naturalistic interventional study. BMC psychiatry. 2018 Dec;18(1):1-9.

Roberts C, Davies J, Maggs RG. Structured community activity for forensic mental health: a feasibility study. Journal of Forensic Practice. 2015 Aug 10.

Robson D, Haddad M, Gray R, Gournay K. Mental health nursing and physical health care: A cross-sectional study of nurses' attitudes, practice, and perceived training needs for the physical health care of people with severe mental illness. International Journal of Mental Health Nursing. 2013 Oct;22(5):409-17.

Rosenbaum S, Sherrington C, Tiedemann A. Exercise augmentation compared with usual care for post-traumatic stress disorder: A randomized controlled trial. Acta Psychiatrica Scandinavica. 2015 May;131(5):350-9.

Roy A, Govindan R, Muralidharan K. The impact of an add-on video assisted structured aerobic exercise module on mood and somatic symptoms among women with depressive disorders: Study from a tertiary care centre in India. Asian Journal of Psychiatry. 2018 Feb 1;32:118-22.

Safdar N, Marryam M, Sheraz S, Malik AN, Amjad I. Effect of supervised exercise on sleep deprivation and quality of life in patients with depression. Rawal Medical Journal. 2019 Feb 15;44(1):113-6.

Sailer P, Wieber F, Pröpster K, Stoewer S, Nischk D, Volk F, Odenwald M. A brief intervention to improve exercising in patients with schizophrenia: a controlled pilot study with mental contrasting and implementation intentions (MCII). BMC psychiatry. 2015 Dec;15:1-2.

Salehi I, Hosseini SM, Haghighi M, Jahangard L, Bajoghli H, Gerber M, Pühse U, Holsboer-Trachsler E, Brand S. Electroconvulsive therapy (ECT) and aerobic exercise training (AET) increased plasma BDNF and ameliorated depressive symptoms in patients suffering from major depressive disorder. Journal of psychiatric research. 2016 May 1;76:1-8.

Savage P, Long C, Hall L, Mackenzie R, Martin L. Reaping the rewards of better fitness. mental health practice. 2009 Feb 10;12(5).

Schneider BC, Moritz S, Hottenrott B, Reimer J, Andreou C, Jelinek L. Association Splitting: A randomized controlled trial of a new method to reduce craving among inpatients with alcohol dependence. Psychiatry research. 2016 Apr 30;238:310-7.

Schuch FB, Vasconcelos-Moreno MP, Borowsky C, Fleck MP. Exercise and severe depression: preliminary results of an add-on study. Journal of affective disorders. 2011 Oct 1;133(3):615-8.

Schuch FB, Vasconcelos-Moreno MP, Borowsky C, Zimmermann AB, Wollenhaupt-Aguiar B, Ferrari P, de Almeida Fleck MP. The effects of exercise on oxidative stress (TBARS) and BDNF in severely depressed inpatients. European archives of psychiatry and clinical neuroscience. 2014 Oct;264:605-13.

Schuch FB, Vasconcelos-Moreno MP, Borowsky C, Zimmermann AB, Rocha NS, Fleck MP. Exercise and severe major depression: effect on symptom severity and quality of life at discharge in an inpatient cohort. Journal of psychiatric research. 2015 Feb 1;61:25-32.

Schulze T, Hahn E, Hahne I, Bergmann N, Fuchs LM, Mähler F, Zierhut MM, Ta TM, Pijnenborg GH, Böge K. Yoga-based group intervention for in-patients with schizophrenia spectrum disorders—a qualitative approach. Frontiers in Psychiatry. 2021 Aug 13;12:715670.

Şenormancı G, Korkmaz N, Şenormancı Ö, Uğur S, Topsaç M, Gültekin O. Effects of exercise on resilience, insight and functionality in patients with chronic schizophrenia in a psychiatric nursing home setting: a randomized controlled trial. Issues in Mental Health Nursing. 2021 Jul 3;42(7):690-8.

Shachar-Malach T, Kazaz RC, Constantini N, Lerer B. Effectiveness of aerobic exercise as an augmentation therapy for inpatients with major depressive disorder: a preliminary randomized controlled trial. Israel Journal of Psychiatry. 2015 Jul 1;52(3):65.

Shimada T, Ito S, Makabe A, Yamanushi A, Takenaka A, Kobayashi M. Aerobic exercise and cognitive functioning in schizophrenia: A pilot randomized controlled trial. Psychiatry Research. 2019 Dec 1;282:112638.

Shimada T, Ito S, Makabe A, Yamanushi A, Takenaka A, Kawano K, Kobayashi M. Aerobic exercise and cognitive functioning in schizophrenia: results of a 1-year follow-up from a randomized controlled trial. Psychiatry Research. 2020 Apr 1;286:112854.

Sistig B, Friedman SH, McKenna B, Consedine NS. Mindful yoga as an adjunct treatment for forensic inpatients: a preliminary evaluation. The Journal of Forensic Psychiatry & Psychology. 2015 Nov 2;26(6):824-46.

Song BK, Kim YS, Kim HS, Oh JW, Lee O, Kim JS. Combined exercise improves gastrointestinal motility in psychiatric in patients. World Journal of Clinical Cases. 2018 Aug 8;6(8):207.

Sørensen M, Bentzen M, Farholm A. Lessons learned from a physical activity intervention in psychiatric treatment: Patient, staff, and leader perspectives. Frontiers in psychiatry. 2020 Mar 2;11:87.

Sørensen M, Bentzen M, Farholm A. Motivational physical activity intervention for psychiatric inpatients: A two phased single-cases experimental study. European Journal of Adapted Physical Activity 2021, 12, 14; doi: 10.5507/euj.2021.008.

Spinelli C, Paradis-Gagné E, Per M, Fleischmann MH, Manova V, Wallace A, Khoury B. Evaluating the use of mindfulness and yoga training on forensic inpatients: a pilot study. Frontiers in psychiatry. 2020 Dec 10;11:614409.

Srivastava S, Bhatia MS, Gautam P, Saha R, Chauhan J. Yoga and exercise intervention study in psychiatry inpatients from a tertiary care teaching hospital. Delhi Psychiatry J. 2015;18(1):138-41.

Stanley SH, Ng SM, Laugharne JD. The ‘Fit for Life’ exercise programme: improving the physical health of people with a mental illness. Psychology, Health & Medicine. 2019 Feb 7;24(2):187-92.

Stanton R, Reaburn P, Happell B. The Effect of Acute Exercise on Affect and Arousal in Inpatient Mental Health Consumers. J Nerv Ment Dis. 2016 Sep;204(9):658-64. doi: 10.1097/NMD.0000000000000510.

Stanton R, Donohue T, Garnon M, Happell B. Participation in and Satisfaction With an Exercise Program for Inpatient Mental Health Consumers. Perspect Psychiatr Care. 2016 Jan;52(1):62-7. doi: 10.1111/ppc.12108.

Stubbs B, Cooper-Evans MS, Durran M, Montenegro JM. An exploration of walking activity among older psychiatric inpatients. International Journal of Therapy and Rehabilitation. 2007 Dec;14(12):532-7.

Tague DB. The effect of improvisational group drumming versus general music therapy versus activity therapy on mood, session behaviors and transfer behaviors of in-patient psychiatric individuals. The Florida State University; 2012.

Tanaka C, Yotsumoto K, Tatsumi E, Sasada T, Taira M, Tanaka K, Maeda K, Hashimoto T. Improvement of functional independence of patients with acute schizophrenia through early occupational therapy: a pilot quasi-experimental controlled study. Clinical Rehabilitation. 2014 Aug;28(8):740-7.

Tazesh B, Shahi MH, Mokri A, Naghavi H, Sharafi SE, Mirsepassi Z. Service Development and Implementation of an Exercise Therapy Unit in a Psychiatric Hospital in Iran: A Brief Report. Iranian Journal of Psychiatry and Behavioral Sciences. 2021 Sep 30;15(3).

Tetlie T, Eik-Nes N, Tom Palmstierna MD. The effect of exercise on psychological & physical health outcomes: preliminary results from a Norwegian forensic hospital. Journal of Psychosocial Nursing & Mental Health Services. 2008 Jul 1;46(7):38.

Tetlie T, Heimsnes MC, Almvik R. Using exercise to treat patients with severe mental illness: how and why? J Psychosoc Nurs Ment Health Serv. 2009 Feb;47(2):32-40. doi: 10.3928/02793695-20090201-14.

Thaller L, Frühauf A, Heimbeck A, Voderholzer U, Kopp M. A comparison of acute effects of climbing therapy with nordic walking for inpatient adults with mental health disorder: A clinical pilot trial. International Journal of Environmental Research and Public Health. 2022 Jun 1;19(11):6767.

Tomasi D, Gates S, Reyns E. Positive Patient Response to a Structured Exercise Program Delivered in Inpatient Psychiatry. Glob Adv Health Med. 2019 May 21;8:2164956119848657. doi: 10.1177/2164956119848657.

Torelly GA, dos Santos Novak P, Bristot G, Schuch FB, de Almeida Fleck MP. Acute effects of mind-body practices and exercise in depressed inpatients: A randomized clinical trial. Mental Health and Physical Activity. 2022 Oct 1;23:100479.

Ujike S, Yasuhara Y, Osaka K, Sato M, Catangui E, Edo S, Takigawa E, Mifune Y, Tanioka T, Mifune K. Encounter of Pepper-CPGE for the elderly and patients with schizophrenia: an innovative strategy to improve patient's recreation, rehabilitation, and communication. The Journal of Medical Investigation. 2019 Feb 15;66(1.2):50-3.

Vancampfort D, De Hert M, Knapen J, Wampers M, Demunter H, Deckx S, Maurissen K, Probst M. State anxiety, psychological stress and positive well-being responses to yoga and aerobic exercise in people with schizophrenia: a pilot study. Disability and rehabilitation. 2011 Jan 1;33(8):684-9.

Visceglia E, Lewis S. Yoga therapy as an adjunctive treatment for schizophrenia: a randomized, controlled pilot study. J Altern Complement Med. 2011 Jul;17(7):601-7. doi: 10.1089/acm.2010.0075. PMID: 21711202.

Wang J, Li Z. Effect of physical exercise on medical rehabilitation treatment of depression. Revista Brasileira de Medicina do Esporte. 2022 Feb 28;28:174-6.

Warren KR, Ball MP, Feldman S, Liu F, McMahon RP, Kelly DL. Exercise program adherence using a 5-kilometer (5K) event as an achievable goal in people with schizophrenia. Biol Res Nurs. 2011 Oct;13(4):383-90. doi: 10.1177/1099800410393272.

Woodward ML, Gicas KM, Warburton DE, White RF, Rauscher A, Leonova O, Su W, Smith GN, Thornton AE, Vertinsky AT, Phillips AA, Goghari VM, Honer WG, Lang DJ. Hippocampal volume and vasculature before and after exercise in treatment-resistant schizophrenia. Schizophr Res. 2018 Dec;202:158-165. doi: 10.1016/j.schres.2018.06.054.

Wright A, Cattan M. Physical activity and the management of depression. Working with Older People. 2009 Apr 13;13(1):15-8.

Wu MK, Wang CK, Bai YM, Huang CY, Lee SD. Outcomes of obese, clozapine-treated inpatients with schizophrenia placed on a six-month diet and physical activity program. Psychiatr Serv. 2007 Apr;58(4):544-50. doi: 10.1176/ps.2007.58.4.544.

Wynaden D, Barr L, Omari O, Fulton A. Evaluation of service users' experiences of participating in an exercise programme at the Western Australian State Forensic Mental Health Services. International journal of mental health nursing. 2012 Jun;21(3):229-35.
